# Tongue swab *Mycobacterium tuberculosis* qPCR for community screening of asymptomatic TB vs. clinic-based triage of symptomatic TB

**DOI:** 10.64898/2026.02.12.26346160

**Authors:** Rachel C. Wood, Alaina M. Olson, Katherine A. Lochner, Rane B. Dragovich, Alexey Ball, Amy Steadman, Tahlia Perumal, Simon C. Mendelsohn, Humphrey Mulenga, Michele Tameris, Denis Awany, Tumelo Moloantoa, Stephanus T. Malherbe, Austin Katona, Fernanda Maruri, Kris M. Weigel, Firdows Noor, Ravindre Panchia, Khuthadzo Hlongwane, Kim Stanley, Yuri F. van der Heijden, Katie Hadley, Gerhard Walzl, Thomas J. Scriba, Neil A. Martinson, Keertan Dheda, Al Leslie, Bernard Fourie, Timothy R. Sterling, Gerard A. Cangelosi, Mark Hatherill, the RePORT South Africa Study Team

## Abstract

**BACKGROUND:** Diagnostic performance of tongue swab *Mycobacterium tuberculosis* PCR has been evaluated for facility-based triage of symptomatic tuberculosis (TB). It is unknown whether tongue swab performance differs for detection of asymptomatic TB in community-based screening.

**METHODS:** Tongue swabs were collected from adult household contacts of TB patients (HHC Cohort), and symptomatic adults presenting to clinics with presumptive TB (Clinic Cohort), at eight South African sites. TB Cases were defined by positive sputum Xpert Ultra or liquid culture, performed in all participants; and matched ∼1:3 (HHC Cohort) or ∼1:2 (Clinic Cohort) to Controls without TB. Tongue swabs in both cohorts were tested by high-volume qPCR; and in the Clinic Cohort, also by sequence-specific magnetic capture (SSMaC) with qPCR.

**RESULTS:** The Clinic Cohort included 217 TB Cases (100% symptomatic) and 437 Controls. The HHC Cohort included 44 TB Cases (84.1% asymptomatic) and 136 Controls. In the Clinic Cohort, sensitivity of SSMaC with qPCR was 73.2% (specificity 94.6%), but not significantly higher than high-volume qPCR (63.8%; *p* = 0.14) (specificity 94.4%). Sensitivity of high-volume qPCR in the Clinic Cohort (63.8%) was significantly higher than the HHC Cohort (34.1%; *p* = 0.0007) (specificity 91.9%). Among HHC, high-volume qPCR sensitivity was 35.1% for asymptomatic TB; 52.2% for TB with abnormal CXR; and 100% for TB with High sputum Xpert Ultra grade.

**CONCLUSIONS:** Sensitivity of tongue swab high-volume qPCR for community-based, household screening for asymptomatic TB was low, approximately half that of facility-based triage for symptomatic TB, but increased with radiographic severity and sputum bacillary load.

**Key points:** Sensitivity of tongue swab high-volume qPCR for community tuberculosis screening among primarily asymptomatic household contacts was low and approximately half that of facility-based triage for symptomatic tuberculosis. Sensitivity was lowest in individuals with normal chest radiography and low bacillary burden.

## Introduction

There were 8.2 million tuberculosis (TB) disease cases reported in 2023, yet an estimated 2.7 million went undetected (1). Barriers to care, stigma, and under-utilization of testing services in people with asymptomatic TB—who do not have or do not recognize symptoms—drive missed diagnoses (2). In high-burden settings, asymptomatic disease may constitute 50–60% of prevalent TB (3); and ∼67% of transmission (4). Absence of symptoms, variable lung pathology and chest radiograph (CXR) severity, and inconsistent detection of *Mycobacterium tuberculosis* (MTB) bacilli or DNA in sputum (3), may contribute to sub-optimal performance of clinical, radiographic and molecular TB screening strategies.

Sputum production poses an infection control hazard and may be difficult to collect from asymptomatic individuals. Tongue swabs are easier, safer, and faster to collect. Previous diagnostic accuracy studies among symptomatic, care-seeking patients (facility-based triage), reported tongue swab sensitivity of 66-81% with Xpert Ultra (5–7); and >90% with manual laboratory processing methods and qPCR (8–11). In a recent multi-site study, tongue swabs assayed using a near-point-of-care platform achieved 85.7% sensitivity and 100% specificity versus a sputum microbiological reference standard (MRS) (12). Modeling suggests that non-invasive sampling, such as tongue swabs, could increase TB case-finding yield (2); and these potential gains are reflected in WHO target product profiles (TPP) for new TB diagnostics (13).

Controlling the global TB epidemic may ultimately require mass community screening, in addition to passive, facility-based triage of symptomatic people with presumptive TB. Community-based risk groups, including household contacts (HHC) of TB patients, people living with HIV (PLHIV), and those with prior TB, are priority candidates for targeted universal TB testing, which might detect disease before symptom onset and limit the duration of transmission (14). However, diagnostic accuracy of tongue swab PCR for community TB screening, particularly for asymptomatic TB, is untested. Resolving this critical evidence gap is essential before tongue swab sampling can be included in policy recommendations for community-based TB screening.

The RePORT South Africa network has established two parallel cohorts, one of symptomatic patients seeking care for presumptive TB; and one of household contacts of TB patients, which allows direct comparison of novel TB screening tests for facility-based triage of symptomatic TB vs. community-based screening for asymptomatic TB(15). We hypothesized that early/mild asymptomatic disease and lower MTB bacillary load might lower tongue swab sensitivity. Since achieving optimal sensitivity is paramount for screening paucibacillary populations, we used laboratory-based, manual tongue swab testing methods: the high-volume qPCR method of Steadman *et al.* 2024 (8) and, to further improve sensitivity, the sequence-specific magnetic capture (SSMaC) with qPCR method of Olson *et al.* 2025 (9).

## Materials & Methods

### Population and Setting

The RePORT South Africa protocol enrolled participants at 8 sites in geographically diverse locations in South Africa (HHC Cohort 3 sites; Clinic Cohort 5 sites). The Clinic Cohort included adults (≥18 years), living with or without HIV, who presented to fixed or mobile healthcare clinics with symptoms consistent with TB. The HHC Cohort included adult household contacts of index TB patients identified by the health care system(15). Eligible HHC had significant exposure within the past 6 months to an adult with untreated/inadequately treated TB, defined as sleeping in the same household or four or more hours of other close exposure per week. All participants provided written informed consent. The protocol was approved by the institutional ethics committee of the participating sites.

### Study Procedures

Universal TB screening was conducted at enrolment for all participants, including symptom screening for any symptom consistent with TB, including persistent (>2 weeks) unexplained cough, fever, weight loss, pleuritic chest pain, night sweats, or any hemoptysis; CXR, reported by a single investigator at each site; and one spontaneous, expectorated sputum sample for Xpert® MTB/RIF Ultra (Cepheid, CA, USA), liquid culture (Mycobacteria Growth Indicator Tube [MGIT], BACTEC, Becton Dickinson, NJ, USA), and acid-fast smear microscopy. The MRS Case definition for microbiologically-confirmed pulmonary TB disease was at least one sputum positive for MTB by MGIT culture and/or Xpert Ultra (excluding trace). Symptomatic and asymptomatic TB were defined as microbiologically-confirmed TB with or without self-reported symptoms of any duration, respectively. Participants without positive sputum MGIT culture or Xpert Ultra were defined as Controls. Spontaneous expectorated sputum collection was attempted for all participants; sputum sample volume and quality were not recorded. Sputum induction was not performed. Each participant provided one tongue swab sample using Regular FLOQSwabs® flocked swabs (520CS01, Copan Italia, Brescia, Italy) prior to sputum collection at their baseline visit. Each swab was stored dry and frozen at −80 °C.

### Sample Selection and Analysis

All tongue swab sample analyses were performed blind to TB status. Laboratory staff were not blinded to study cohort. Tongue swab samples from the HHC Cohort, which completed enrollment earlier, were tested prior to those from the Clinic Cohort. Tongue swab samples were tested from TB Cases and matched Controls in a ratio of approximately 1:2 in the Clinic Cohort; and approximately 1:3 in the HHC Cohort. In both cohorts, propensity score was used to rank potential Controls by sex, age, HIV status, and enrollment site. These variables were selected *a priori* based on known associations with TB susceptibility and disease presentation.

All HHC Cohort samples were tested using the high-volume qPCR protocol (8) with adaptations for frozen samples. Protocol details can be found in the **Supplemental Methods**.

All Clinic Cohort tongue swab samples were allocated 1:1 to either high-volume qPCR or SSMaC with qPCR. High-volume qPCR followed the same method as for HHC Cohort samples. Minor lab-specific instrumentation differences are described in **Supplemental Methods**.

The remaining samples were tested by SSMaC with qPCR, IS*6110-* and IS*1081*-targeted probe-based DNA capture, and qPCR (9); details in **Supplemental Methods**.

### Data Analysis

Analyses and data visualization were performed in GraphPad Prism, MedCalc, and Microsoft Excel. Sensitivity and specificity, along with associated 95% binomial confidence intervals (95% CI), were calculated for discrimination of TB Cases and Controls in the HHC and Clinic Cohorts; and between tongue swab testing methods. The “exact” Clopper-Pearson interval method was used to estimate 95% CIs. Differences in proportions were tested with two-sided Z-tests for two independent populations (α = 0.05).

## Results

Tongue swab samples were available for 44 TB Cases and 136 matched Controls in the HHC Cohort (N = 180). In the Clinic Cohort, 446 Controls were matched to 227 TB Cases with available tongue swabs (N = 673). Sputum samples were available for all participants in both cohorts.

Two sets of samples processed and tested by SSMaC with qPCR showed amplification in the negative control; upon rerun one set resolved (four positive swab results retained) and one did not (two positive swabs excluded). Therefore, following exclusion of these samples, 217 TB Cases and 437 Controls in the Clinic Cohort (N = 654) were included in analyses (**Figure 1**). For samples tested by high-volume qPCR, results based on the fluorescence of IS*1081* were not used, because the signals were weaker than those for IS*6110*.

**Figure 1.**
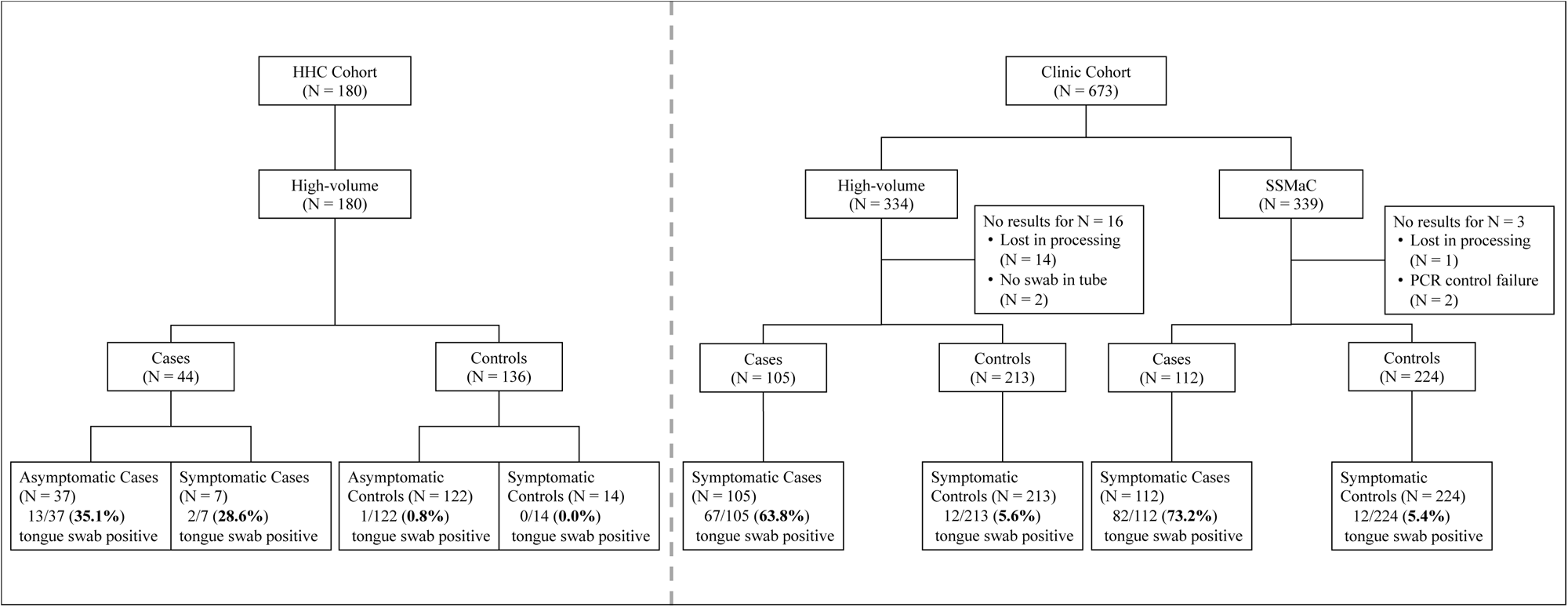
Participant TB Case/Control status, symptom status and tongue swab qPCR results in the HHC and Clinic cohorts. HHC, household contact. SSMaC, sequence-specific magnetic capture.

There were 159/180 (88.3%) asymptomatic participants in the HHC Cohort, including 37/44 (84.1%) TB Cases. All (100%) participants in the Clinic Cohort were symptomatic. Demographic and clinical characteristics of participants in the HHC and Clinic Cohorts are shown in **Tables 1** and **2**, respectively. The Clinic Cohort is differentiated by analytical method in **Table S2.**

**Table 1.**
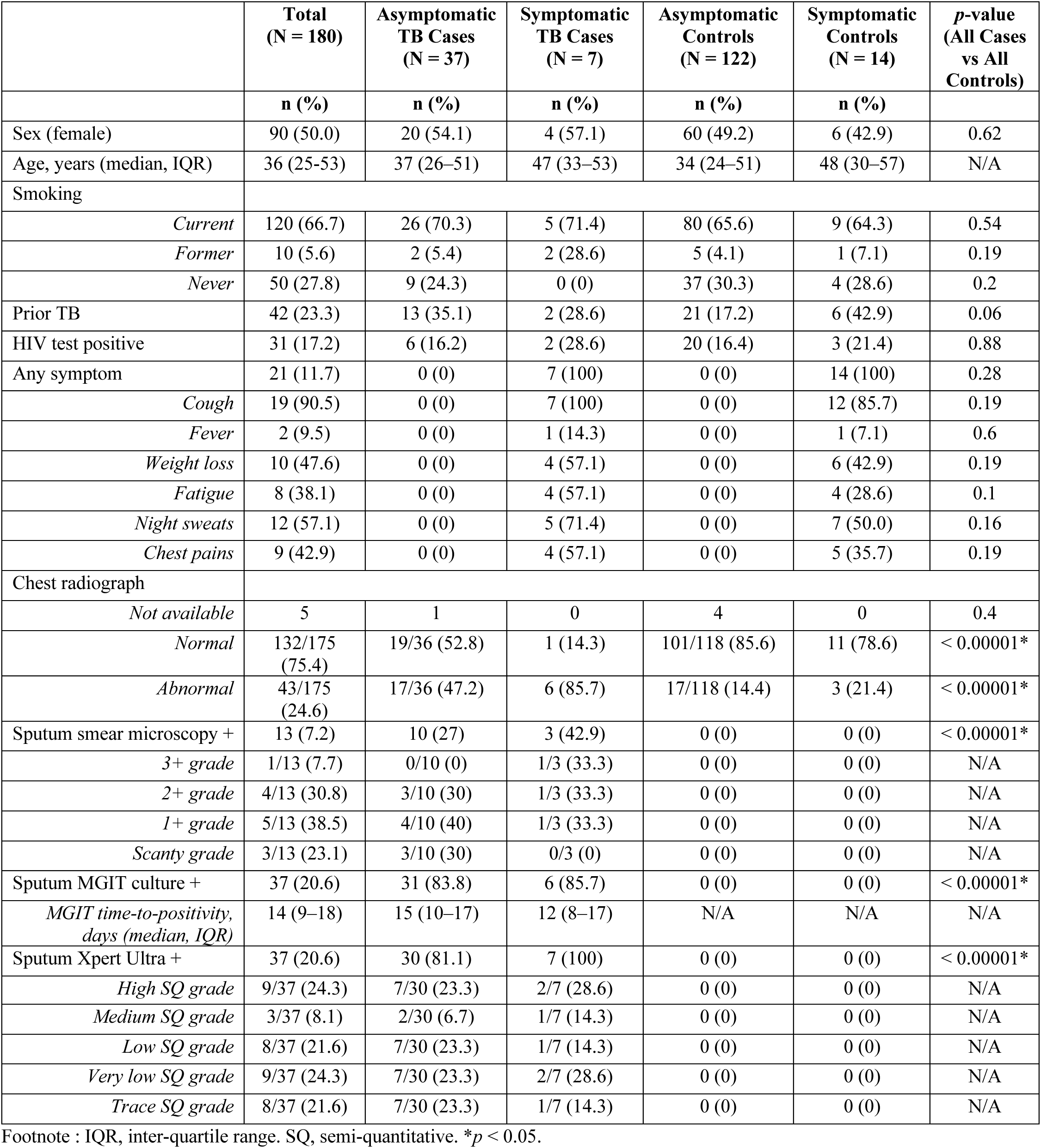
Demographic and clinical characteristics of participants in the HHC Cohort.

**Table 2.** Demographic and clinical characteristics of participants in the Clinic Cohort.

|  | **Total**<br>**(N = 673)** | **Symptomatic TB Cases**<br>**(N = 227)** | **Symptomatic Controls**<br>**(N = 446)** | **p-value** |
| --- | --- | --- | --- | --- |
|  | **n (%)** | **n (%)** | **n (%)** |  |
| Sex (female) | 245 (36.4) | 80 (35.2) | 165 (37.0) | 0.65 |
| Age, years (median, IQR) | 36 (28–44) | 36 (28–43) | 36 (28–44) | N/A |
| Smoking |  |  |  |  |
| *Current* | 299 (44.4) | 81 (35.7) | 218/445 (49.0) | 0.001* |
| *Former* | 77 (11.4) | 42 (18.5) | 35/445 (7.9) | < 0.00001* |
| *Never* | 296 (44.0) | 104 (45.8) | 192/445 (43.1) | 0.50 |
| Prior TB | 148 (22.0) | 38 (16.7) | 110 (24.7) | 0.02* |
| HIV test positive | 204 (30.3) | 82 (36.1) | 122 (27.4) | 0.02* |
| Any symptom | 673 (100) | 227 (100) | 446 (100) | 0.62 |
| *Cough* | 588 (87.4) | 213 (93.8) | 375 (84.1) | 0.0003* |
| *Fever* | 215 (31.9) | 88 (38.8) | 127 (28.5) | 0.01* |
| *Weight loss* | 365 (54.2) | 165 (72.7) | 200 (44.8) | < 0.00001* |
| *Fatigue* | 323 (48.0) | 141 (62.1) | 182 (40.8) | < 0.00001* |
| *Night sweats* | 370 (55.0) | 149 (65.6) | 221 (49.6) | 0.0001* |
| *Chest pains* | 258 (38.3) | 111 (48.9) | 147 (33.0) | < 0.00001* |
| Chest radiograph |  |  |  |  |
| *Not available* | 332 | 119 | 213 | 0.22 |
| *Normal* | 176/341 (51.6) | 21/108 (19.3) | 155/233 (66.5) | < 0.00001* |
| *Abnormal* | 165/341 (48.4) | 87/108 (80.6) | 78/233 (33.5) | < 0.00001* |
| Sputum smear microscopy + | 165/669 (24.7) | 161/224 (71.9) | 4/445 (0.9) | < 0.00001* |
| *Not done* | 4 | 3 | 1 | 0.07 |
| *3+ grade* | 45/165 (27.3) | 45/161 (28.0) | 0/4 (0.0) | 0.22 |
| *2+ grade* | 53/165 (32.1) | 53/161 (32.9) | 0/4 (0.0) | 0.16 |
| *1+ grade* | 38/165 (23.0) | 38/161 (23.6) | 0/4 (0.0) | 0.27 |
| *Scanty grade* | 28/165 (17.0) | 24/161 (14.9) | 4/4 (100) | < 0.00001* |
| Sputum MGIT culture + | 206/668 (30.8) | 206/226 (91.2) | 0/442 (0.0) | < 0.00001* |
| *Not done* | 5 | 1 | 4 | 0.55 |
| *MGIT time-to-positivity, days*<br>*(Median, IQR)* | 9 (6–14) | 9 (6–14) | N/A | N/A |
| Sputum Xpert Ultra + | 221/669 (33.0) | 214/226 (94.7) | 7/443 (1.6) | < 0.00001* |
| *Not done* | 4 | 1 | 3 | 0.73 |
| *High SQ grade* | 77/221 (34.8) | 77/214 (36.0) | 0/7 (0.0) | 0.05* |
| *Medium SQ grade* | 42/221 (19.0) | 42/214 (19.6) | 0/7 (0.0) | 0.2 |
| *Low SQ grade* | 56/221 (25.3) | 56/214 (26.2) | 0/7 (0.0) | 0.12 |
| *Very low SQ grade* | 26/221 (11.8) | 26/214 (12.1) | 0/7 (0.0) | 0.33 |
| *Trace SQ grade* | 19/221 (8.6) | 12/214 (5.6) | 7/7 (100) | < 0.00001* |
| *SQ grade not recorded* | 1/221 (0.5) | 1/214 (0.4) | 0/7 (0.0) | 0.87 |

Footnote : SQ, semi-quantitative. IQR, inter-quartile range. \* $p < 0.05$ .

### Sensitivity and Specificity of Tongue Swab qPCR by Cohort

Sensitivity of SSMaC with qPCR was 73.2%; but was not significantly higher than the high-volume qPCR method in the Clinic Cohort (*p* = 0.14). The high-volume qPCR method was significantly more sensitive in the Clinic Cohort (63.8%, 95% CI 53.9–73.0) than the HHC Cohort (34.1%, 95% CI 20.5–49.9; *p* = 0.0007; **Table 3**).

**Table 3.** Sensitivity and specificity of tongue swab qPCR in the Clinic and HHC Cohorts, stratified by symptom status.

|  | Sensitivity |  | Specificity |  |
| --- | --- | --- | --- | --- |
|  | n/N | % (95% CI) | n/N | % (95% CI) |
| **Clinic Cohort** |  |  |  |  |
| High-volume qPCR | 67/105 | 63.8<br>(53.9–73.0) | 201/213 | 94.4<br>(90.4–97.1) |
| SSMaC with qPCR | 82/112 | 73.2<br>(64.0–81.1) | 212/224 | 94.6<br>(90.8–97.2) |
| **HHC Cohort** |  |  |  |  |
| High-volume qPCR | 15/44 | 34.1<br>(20.5–49.9) | 125/136 | 91.9<br>(86.0–96.0) |
| *Symptomatic* | 2/7 | 28.6<br>(3.67–71.0) | 14/14 | 100<br>(76.84–100) |
| *Asymptomatic* | 13/37 | 35.1<br>(20.21–52.5) | 111/122 | 91.0<br>(84.44–95.4) |

Footnote: SSMaC, sequence-specific magnetic capture. HHC, household contact.

Several samples in each cohort were collected with an alternative flocked swab with a smaller head than the Copan FLOQSwab. In the HHC Cohort, 74% were Copan swabs (61% Cases, 78% Controls); and in the Clinic Cohort, 92% of swab samples were Copan swabs. (93% Cases, 91% Controls) Notably, swab type did not affect overall sensitivity **(Table S3).**

Tongue swab high-volume qPCR was false positive in 1/136 (0.7%) HHC Cohort Control participants (no prior TB reported); and 12/213 (5.6%) Clinic Cohort Control participants, with 2/12 (16.7%) reporting prior TB and 1/12 (8.3%) with sputum trace Xpert Ultra and scanty smear positive. SSMaC with qPCR was false positive in 12/224 (5.4%) Clinic Cohort Control participants, with 4/12 (33.3%) reporting prior TB.

### Severity of Disease and Bacillary Load

Within the HHC Cohort, tongue swab high-volume qPCR sensitivity was not significantly different between symptomatic (28.6%; 2/7) versus asymptomatic (35.1%; 13/37) TB Cases (*p* = 0.74), though the number of symptomatic TB Cases was too small to draw definitive conclusions (**Table 3**). Tongue swabs were significantly more sensitive among TB Cases with an abnormal (52.2%, 95% CI 30.6–73.2) than with a normal CXR (15.0%, 95% CI 3.2–37.9; *p* = 0.011); and tended to be more sensitive among those who had previous TB (53.3%, 95% CI 26.6–78.7) versus without previous TB (24.1%, 95% CI 10.3–43.5; *p* = 0.052).

Test positivity of tongue swab qPCR, sputum Xpert Ultra, and sputum smear was higher in the Clinic Cohort (64%, 95%, 72%, respectively) than the HHC Cohort (34%, 80%, 30%; *p* = 0.001, *p* = 0.002, *p* < 0.0001, respectively) (**Figure 2**). Tongue swab and sputum smear test positivity among asymptomatic participants were also higher among those with High and Medium sputum Xpert Ultra semi-quantitative grades (100%, 89%, respectively) than those with paucibacillary sputum (Low, Very Low, and Trace sputum Xpert Ultra semi-quantitative grades) (19%, 10%; *p* < 0.0001, *p* < 0.0001, respectively) (**Figure S1**).

**Figure 2.**
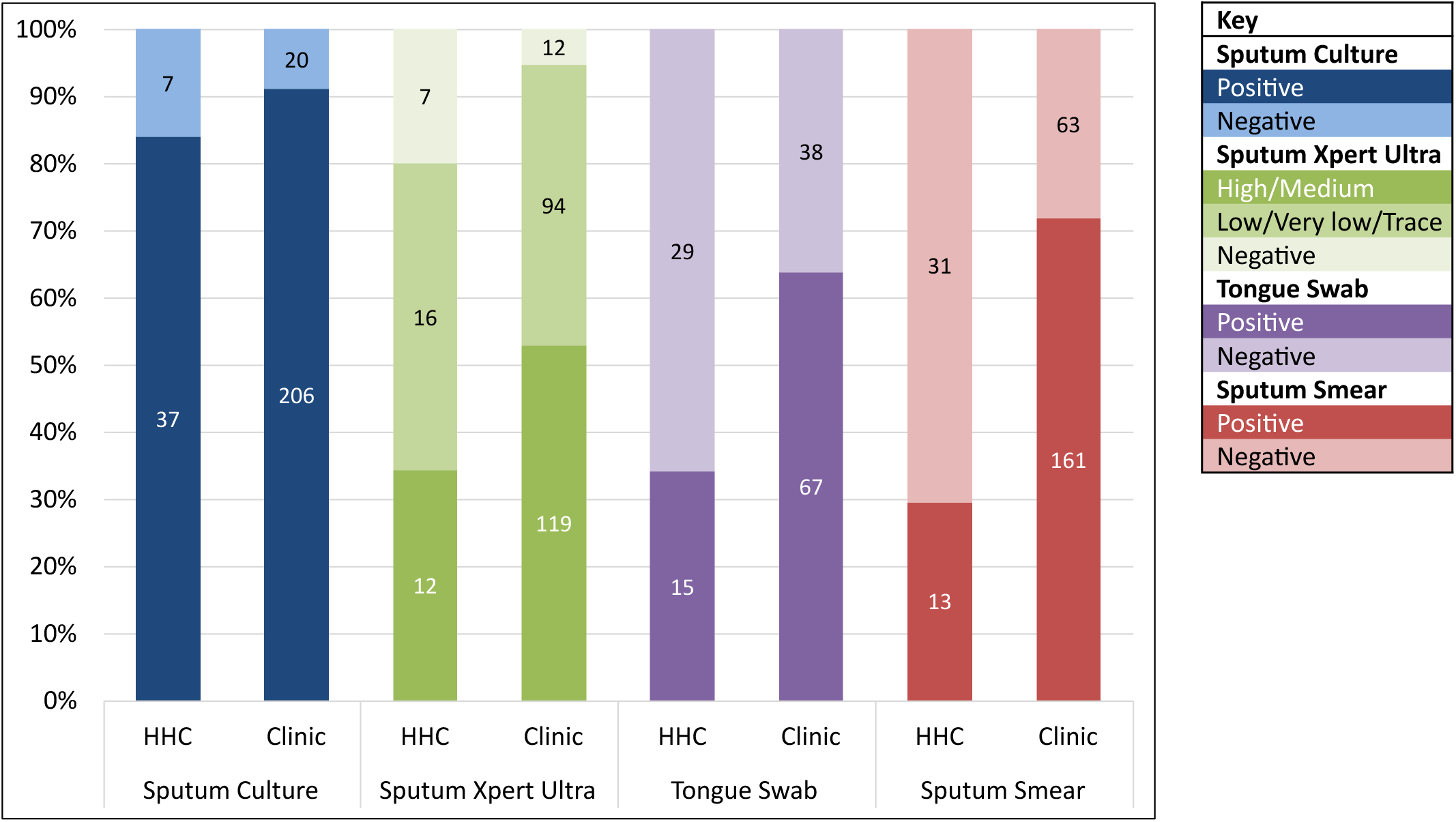
Comparison of frequencies (%) of sputum culture, sputum Xpert Ultra, tongue swab (high-volume qPCR method), and sputum smear positivity versus negativity in the HHC and Clinic Cohorts. Positive sputum Xpert Ultra results are divided into High/Medium and Low/Very low/Trace categories. The numbers within the bars are the test result N. Note that test categories are arranged in descending order of positivity among HHC, which is not the same in the Clinic Cohort, where sputum Xpert Ultra had a higher positivity than culture, and smear had a higher positivity than tongue swabs. Test categories are presented in ascending order of the difference in positivity rates between cohorts by test, which were 7.1%, 14.7%, 29.7%, and 42.3%, respectively. HHC, household contact.

Consistent with higher sputum bacillary load, TB Cases in the Clinic Cohort showed a higher frequency of High and Medium Xpert Ultra grades (52.7% vs 27.3%; *p* = 0.015), and shorter sputum culture time-to-positivity (median 9 vs 14 days; *p* = 0.008, two-tailed Mann-Whitney U test), compared to TB Cases in the HHC Cohort (**Figure S2**). A similar proportion of symptomatic TB Cases in the Clinic and HHC Cohorts had an abnormal CXR (80.6% vs 85.7%), but only 47.2% of asymptomatic TB Cases in the HHC Cohort had an abnormal CXR.

### Sensitivity of Tongue Swab qPCR by Sputum Bacillary Load

When stratified by sputum Xpert Ultra semi-quantitative grade, sensitivity of tongue swab high-volume qPCR was highest (100%) in HHC Cohort TB Cases with High semi-quantitative grade; and was similar between the HHC and Clinic Cohorts for TB Cases with sputum High or Medium semi-quantitative grades (**Table 4**). However, sensitivity in the HHC Cohort fell to less than half that of the Clinic Cohort for TB Cases with a sputum Low semi-quantitative grade.

**Table 4.** Sensitivity of tongue swab analytic method by cohort and sputum Xpert Ultra semi-quantitative grade.

|  | **HHC Cohort** |  | **Clinic Cohort** |  |  |  |
| --- | --- | --- | --- | --- | --- | --- |
|  | **High-volume qPCR** |  | **High-volume qPCR** |  | **SSMaC with qPCR** |  |
|  | **n/N** | **% (95% CI)** | **n/N** | **% (95% CI)** | **n/N** | **% (95% CI)** |
| Sputum Xpert Ultra grade |  |  |  |  |  |  |
| High | 9/9 | 100 (66.4, 100) | 27/31 | 87.1 (70.2, 96.4) | 42/43 | 97.7 (87.7, 100) |
| Medium | 2/3 | 66.7 (9.4, 99.2) | 16/19 | 84.2 (60.4, 96.6) | 21/21 | 100 (83.9, 100) |
| Low | 2/8 | 25 (3.2, 65.1) | 16/29 | 55.2 (35.7, 73.6) | 15/24 | 62.5 (40.6, 81.2) |
| Very low | 1/9 | 11.1 (0.3, 48.3) | 5/14 | 35.7 (12.8, 64.9) | 2/11 | 18.2 (2.3, 51.8) |
| Trace | 1/8 | 12.5 (0.3, 52.7) | 1/5 | 20.0 (0.5, 71.6) | 1/6 | 16.7 (0.4, 64.1) |
| Sputum culture+ / Xpert– | 0/7 | 0 (0.0, 41.0) | 2/7 | 28.6 (3.7, 71.0) | 0/5 | 0 (0.0, 52.2) |
| Excluding sputum Xpert Trace+ result | 14/36 | 38.9 (23.1, 56.5) | 66/100 | 66.0 (55.9, 75.2) | 80/104 | 76.9 (67.6, 84.6) |
| Excluding sputum culture+ / Xpert– | 15/37 | 40.5 (24.8, 57.9) | 65/98 | 66.3 (56.1, 75.6) | 81/105 | 77.1 (67.9, 84.8) |
| Excluding sputum Xpert Trace+ and culture+ / Xpert– | 14/29 | 48.3 (29.5, 67.5) | 64/93 | 68.8 (58.4, 78.0) | 80/99 | 80.8 (71.7, 88.0) |

Footnote: One Xpert Ultra-confirmed TB Case in the Clinic Cohort (tongue swab analyzed by SSMaC with qPCR) did not have a semi-quantitative grade recorded. HHC, household contact. SSMaC, sequence-specific magnetic capture.

In the Clinic Cohort, the SSMaC with qPCR method appeared to improve upon sensitivity compared to the high-volume method for TB Cases with sputum High, Medium, or Low Xpert Ultra semi-quantitative grades, but not for those with sputum Very Low, Trace, or Negative (but culture-positive) Xpert Ultra results. However, the difference in sensitivity between these two methods was not significant at any grade except for Medium (*p* <0.0001).

### Sensitivity of Tongue Swab qPCR in People Living with HIV

Sensitivity of tongue swab high-volume qPCR was similar in people with and without HIV in the HHC Cohort, although there were few TB Cases among PLHIV (n = 8) (**Figure 3**). In the Clinic Cohort, sensitivity of the high-volume qPCR method was lower among PLHIV than those without HIV (*p* = 0.001), and lower among PLHIV tested with high-volume qPCR than those tested by SSMaC with qPCR (*p* = 0.028).

**Figure 3.**
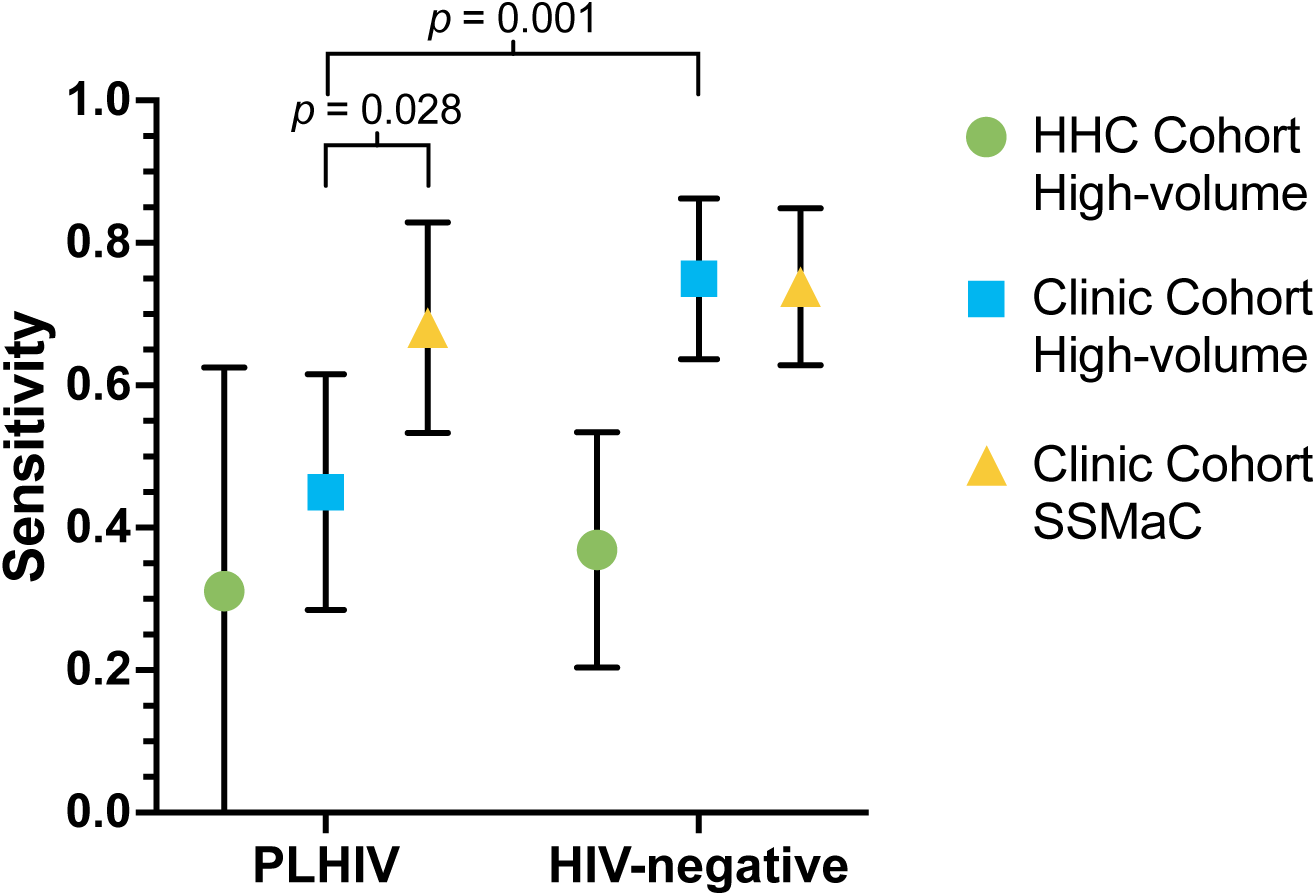
Sensitivity of tongue swab analyses by HIV status as shown by point estimates with error bars indicating 95% confidence intervals. *p*-values shown were significant at *p* < 0.05 by two-population proportion z-test. HHC, household contact. SSMaC, sequence-specific magnetic capture. PLHIV, people living with HIV.

### Evaluation of Tongue Swabs in TB Screening Approaches for HHC

In the HHC Cohort, adding tongue swab high-volume qPCR (sensitivity 34.1%, 95% CI 20.5–49.9) to symptom screening (15.9%, 95% CI 6.6–30.1) marginally increased yield of TB Cases to 45.5% (95% CI 30.4–61.2) (**Figure 4**). Addition of tongue swab sampling to CXR screening (combined sensitivity 59.1%, 95% CI 43.3–73.7) added minimal benefit to CXR alone (sensitivity 53.5%, 95% CI 37.7–68.8).

**Figure 4.**
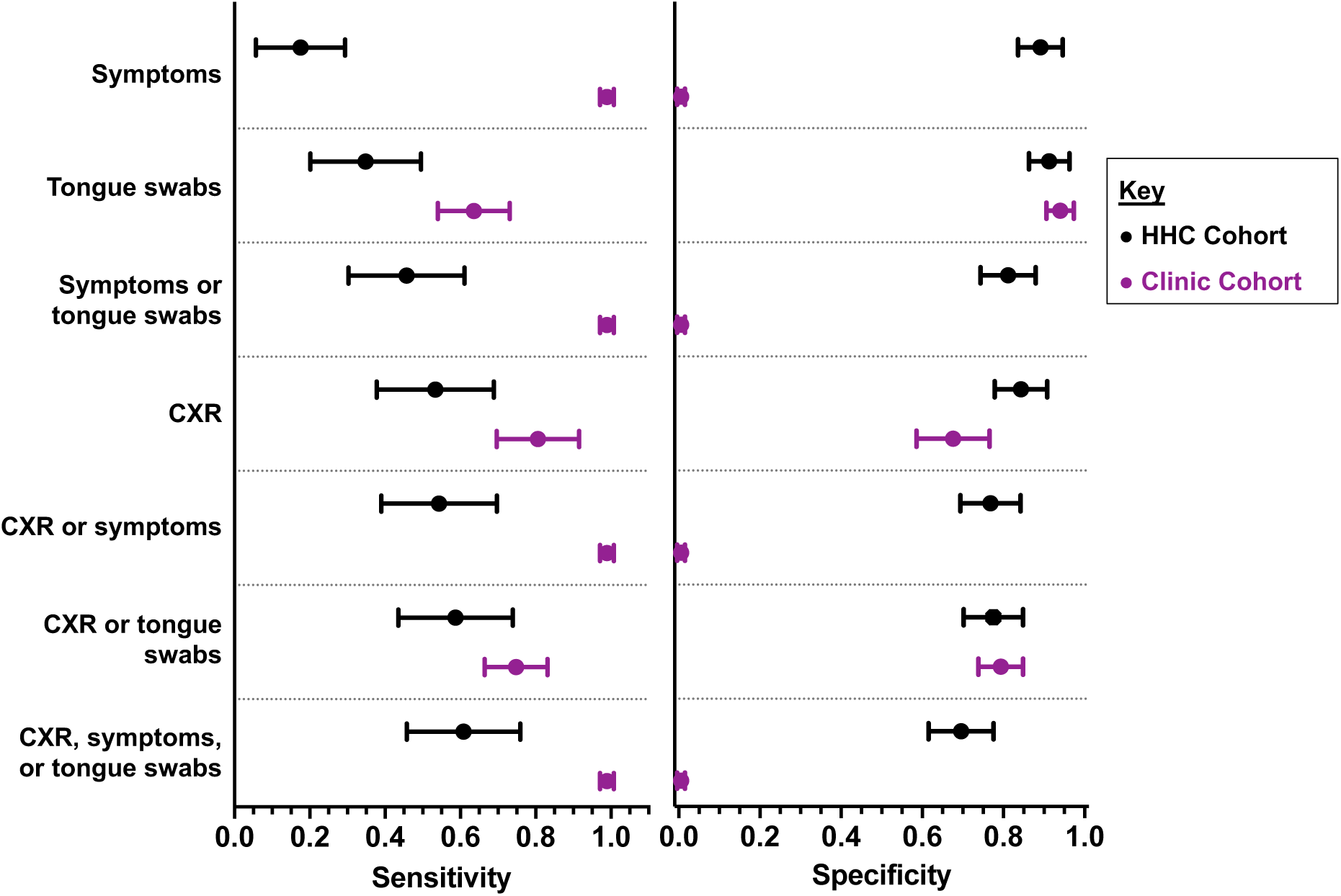
Sensitivities and specificities (point estimates with error bars indicating 95% confidence intervals) of different screening approaches among HHC and Clinic Cohort participants tested by the high-volume qPCR method. CXR, chest radiograph.

A HHC screening strategy combining all three approaches (symptoms, CXR, and tongue swab sampling) achieved a sensitivity of 61.4% (95% CI 45.5–75.6) (specificity 69.9%, 95% CI 61.4–77.4). Specificity was highest for symptom screening and tongue swab testing alone at 89.7% (95% CI 83.3–94.3) and 91.9% (95% CI 86.0–95.9), respectively. However, specificity fell when combined test approaches were considered: symptom or tongue swab (81.6%, 95% CI 74.1–87.7); CXR or tongue swab (77.9%, 95% CI 70.0–84.6); symptom or CXR (77.2%, 95% CI 69.2–84.0); and symptom, CXR, or tongue swab (69.9%, 95% CI 61.4–77.4) (**Figure 4**).

## Discussion

We showed that sensitivity of tongue swab high-volume qPCR for community-based TB screening among predominantly asymptomatic HHC was low; and was approximately half that of facility-based triage among symptomatic clinic attendees with presumptive TB. Sputum Xpert Ultra semi-quantitative grade and CXR findings suggest that this difference in cohort performance likely reflected lower sputum bacillary load and mild/earlier disease among asymptomatic HHC, compared to symptomatic clinic attendees. Consistent with prior facility-based triage studies, tongue swab qPCR was more sensitive among symptomatic individuals with high sputum bacillary load, regardless of the swab testing method (5, 6, 9, 11).

These findings emphasize that community-based TB screening and facility-based triage are distinct use-cases, with different symptom profiles, radiological disease severity, and sputum bacillary burden.

Among symptomatic individuals in the Clinic Cohort, tongue swab qPCR, using both the high-volume and SSMaC methods, closely approached the WHO Target Product Profile (TPP) for a high specificity TB screening test (60% sensitivity; 98% specificity). However, in the HHC Cohort, tongue swab qPCR using the high-volume method fell short of the WHO TPP sensitivity benchmark of 60% (16). These findings, which contrast with favorable reports from triage of symptomatic clinic attendees (5, 6, 9, 11), are consistent with lower impact and cost-effectiveness of tongue swab qPCR for community-based screening of asymptomatic TB. Some community cohorts have reported low rates of spontaneous expectorated sputum collection, which may be an indication for tongue swab sampling (17). All HHC in this study were able to produce a sputum sample without induction; we speculate that sensitivity in sputum-scarce populations might be lower than we have observed.

Improved tongue swab qPCR test sensitivity for paucibacillary and early/mild disease will be essential for efficient community screening of asymptomatic TB. Options include collecting multiple swabs per person; use of foam swabs with higher biomass collection potential (9); and storage in buffer rather than dry freezing. Better performance might also be achieved by use of SSMaC or other new high-sensitivity analytical methods (12, 18). It is not yet known if the necessary gains in sensitivity can be achieved at low sputum MTB bacillary load, where improvement is most crucial. By contrast, tongue swab qPCR sensitivity was excellent (100%) at high bacillary load, even in the HHC Cohort, and comparable to that in the Clinic Cohort. SSMaC with qPCR may be more sensitive than the high-volume qPCR method, especially among PLHIV, in whom SSMaC with qPCR outperformed despite analyzing only ∼20% of the total sample volume (∼50% for high-volume qPCR). Unlike high-volume qPCR, which interrogates crude sample lysate directly, SSMaC concentrates and purifies MTB DNA, which may enhance qPCR performance. Both methods targeted multicopy IS*6110* and IS*1081*; the qPCR protocol used with SSMaC co-amplified the targets in a single channel (same fluorophore), whereas high-volume qPCR used distinct fluorophores. Future work will test SSMaC with qPCR and new high-sensitivity methods (12, 18), in HHC and Clinic cohorts currently being enrolled by the RePORT South Africa network.

To our knowledge, no prior study has reported tongue swab qPCR diagnostic accuracy in a community-based HHC cohort, in which participants were largely asymptomatic. One study evaluated performance in people living with HIV (PLHIV) who were starting antiretroviral therapy; 58% were asymptomatic and tongue swabs showed 22% sensitivity within that subgroup (19), which is comparable to the low sensitivity observed for community-based screening in this study. Tongue swab sensitivity did not differ by symptom status among HHC, but sub-group sizes were small. Among HHC, we found that parallel symptom and CXR screening showed a sensitivity of 55%; adding tongue swab testing modestly increased sensitivity to 61%. If CXR were unavailable, adding tongue swab testing to symptom screening alone significantly improved diagnostic yield (16% vs 46%).

This study had limitations that affect generalizability of the findings. HHC Cohort samples were tested by the high-volume qPCR method, which may be less sensitive than SSMaC with qPCR. Although the same high-volume qPCR assay protocol was used for both cohorts, we cannot exclude the possibility that differences in instrumentation might have affected assay performance. However, given that sensitivity for TB cases with high sputum MTB bacillary load was very similar, it seems more likely that differences in overall sensitivity were due primarily to the predominance of paucibacillary, asymptomatic TB among HHC. Tongue swab sensitivity for community-based screening of asymptomatic TB might improve using a different assay method (12, 18), but only if greater sensitivity can be achieved for pauci-bacillary disease. Ascertainment of TB cases in this study required sputum expectoration and therefore tongue swab qPCR sensitivity in individuals who were sputum-unproductive was not tested. It was surprising that several tongue swab qPCR results in the Clinic Cohort were positive despite a negative sputum MRS, but discordant tongue swab results could conceivably reflect true disease if the expectorated sputum sample was inadequate, or a prior unrecognized TB episode. Finally, the low TB prevalence among HHC limited the statistical power of sub-analyses, for example among PLHIV.

## Conclusion

Sensitivity of tongue swab high-volume qPCR for community-based TB screening among largely asymptomatic HHC was low; and was approximately half that of facility-based triage of symptomatic individuals with presumptive TB. This striking difference in test performance highlights important differences between the screening and triage use-cases and likely reflects the predominance of early/mild, asymptomatic TB disease in community settings. Impact and cost-benefit projections for community-based tongue swab TB screening should be based on data from this same population. Further optimization to improve tongue swab sensitivity for asymptomatic, pauci-bacillary TB is needed to meet WHO performance targets for community-based screening.

## Data Availability

All data produced in the present study are available upon reasonable request to the authors, subject to approval of a concept proposal by the RePORT South Africa network

## Acknowledgements

The authors thank the funders, study staff, study participants, and their families.

## Author contributions

GAC, TRS, and MH conceived the idea, raised funds, and provided the resources. GW, TJS, TRS, and MH wrote the study protocol. GW, TRS, MH, NAM, KD, AL, and BF provided study oversight. TP, SCM, MT, TM, STM, and FN were responsible for field site activities, including recruitment, clinical management, and data collection. KHa provided operational support and project management. HM, DA, AK, FM, RP, KHl, KS, and YFvdH cleaned and verified the underlying data. RCW, AMO, KAL, RBD, AB, AS, KMW, and GAC performed assays and analysed the data. RCW, AMO, GAC, SCM, and MH interpreted the results and wrote the first draft. All authors had full access to the data, confirm the integrity of the data and its presentation, agree with its interpretation as discussed in the manuscript, and reviewed, revised, and approved the manuscript before submission. The corresponding author had final responsibility for the decision to submit for publication.

## Funding statement

This research was supported by the RePORT South Africa network with funds received from CRDF Global (University of Cape Town, G-DAA3-19-66875-1; University of Cape Town Lung Institute, G-DAA9-20-66917-1; Vanderbilt University Medical Center, G-DAA9-20-66870-1; Stellenbosch University, G-DAA9-20-66918-1; Africa Health Research Institute G-202403-71799; University of Pretoria, G-DAA9-20-66880-1 and Wits Health Consortium, G-DAA9-20-66878-1); the US National Institutes of Health (NIH; Stellenbosch University, U01AI152075); and the South African Medical Research Council (SAMRC). The content and findings reported are the sole deduction, view and responsibility of the researcher and do not reflect the official position and sentiments of the SAMRC or the NIH.

## Conflict of Interest Statement

RCW, AMO and GAC report grants from the National Institutes of Health and RePORT South Africa; RCW and GAC report a grant from the Firland Foundation; SCM reports research grants from the Gates Foundation, CRDF Global, the SAMRC and an honorarium from CRDF Global; RP reports grants from the National Institutes of Health; YFvdH and TRS report grants from the National Institutes of Health to Vanderbilt University Medical Center; GW reports grants from the National Institutes of Health and the South African National Research Foundation; TJS reports grants from the Gates Foundation, CRDF Global; SAMRC, Wellcome Trust and Coefficient Giving; NAM reports leadership roles for Wits Health Consortium and Setshaba Research Centre; BF reports in-kind support from Longhorn Vaccines and Diagnostics; GAC reports consulting for Formulatrix and travel and in-kind support from Copan Diagnostics; MH reports research grants from CRDF Global and SAMRC to University of Cape Town. KMW, KAL, RBD, AB, AS, TP, HM, MT, DA, TM, STM, AK, FM, FN, KhH, KS, KaH, KD and AL declare no conflicts of interest.

## Notes

### Author Declarations

The Human Research Ethics Committee of the University of Cape Town gave ethical approval for this work

