## Supplementary Material for "Tongue swab *Mycobacterium tuberculosis* qPCR for community screening of asymptomatic TB vs. clinic-based triage of symptomatic TB"

**Table of Contents**


**Pages 3-5:** **Supplemental Methods**

**Pages 6-7:** **Table S1.** List of RePORT South Africa study team members

**Pages 8-9:** **Table S2.** Demographic and clinical characteristics of participants in the Clinic Cohort, differentiated by testing method

**Pages 10:** **Table S3.** Sensitivity and specificity of tongue swab qPCR in the Clinic and HHC Cohorts

**Page 11:** **Figure S1.** Tongue swab, sputum culture and smear, chest radiograph, and symptom positivity by sputum Xpert Ultra grade among (A) symptomatic and (B) asymptomatic participants

**Page 12:** **Figure S2.** Proportion of sputum Xpert Ultra semi-quantitative grades in TB Cases by cohort.

### Supplemental Methods

#### Household Contact Cohort samples

Household Contact (HHC) Cohort samples were tested at Global Health Labs (Bellevue, WA, USA) by RCW, KAL, and AB. In the published protocol (1), tongue swabs (Regular FLOQSwabs®, 520CS01, Copan Italia, Brescia, Italy) were collected from participants and immediately placed in tubes containing 500 µL 1x Tris-EDTA (TE) buffer and tested the same day. In this study, the dry tongue swabs were removed from the -80 °C freezer and heated at 100 °C for 10 minutes to achieve MTB sterilization, 500 µL TE buffer was then added, and samples were vortexed for 15 seconds at high speed. Samples were then incubated for a further 10 minutes at 100 °C and tested as previously described in Steadman *et al.*, 2024 (1). For the qPCR analysis, 250 µL of the sample was divided into 5 reaction wells, each containing 50 µL to maximize the amount of sample analyzed. One positive control (healthy volunteer tongue swab spiked with an estimated 10,000 *Mycobacterium tuberculosis* H37Ra cells (cultured and stored frozen at pre-determined concentrations) and one negative control (500 µL of TE buffer) were processed and tested alongside every 14 clinical samples. A 10-fold dilution standard curve (0.1 to 100,000 genomes per reaction) using quantitative genomic *Mycobacterium tuberculosis* H37Rv DNA (ATCC 25618DQ, Manassas, Virginia, USA) diluted in molecular grade water was also plated in duplicate. Detection of amplification, with a Cq cut-off threshold of 40 and a fluorescence export threshold of 100,000 Δ Rn for the IS*6110* target sequence, in any of the 5 replicate wells was considered a positive test result. Results for IS*1081* were also obtained using a different fluorescent probe in the multiplex qPCR assay.

**Clinic Cohort Samples**

*High-volume qPCR*

All Clinic Cohort samples were tested at the University of Washington (Seattle, WA, USA). Both the HHC Cohort samples and half of the Clinic Cohort samples were tested using the high-volume qPCR method; however, there were some lab-specific instrumentation differences for the Clinic Cohort: a Fisher Scientific Isotemp heat block (88-860-023, Waltham, MA, USA) rather than a Thermo Scientific heat block (88-870-006, Waltham, MA, USA); a CFX96 Deep Well thermocycler (Bio-Rad 3600037, Hercules, CA, USA) rather than a QuantStudio 5 (Applied Biosystems A34322, Waltham, MA, USA) (qPCR cycling parameters were sustained); and fluorescence thresholds set to 500 RFU (Bio-Rad) versus 100,000 Δ Rn (QuantStudio 5), as described above. The high-volume qPCR testing of the Clinic Cohort samples was done by RBD and RCW.

*SSMaC with qPCR*

The remaining Clinic Cohort samples were tested by SSMaC with qPCR, IS*6110*- and IS*1081*-targeted probe-based DNA capture and qPCR by AMO and RBD (2). Unlike the previously described protocol, which used archived lysates, sample processing began from dry, frozen (-80 °C) tongue swabs. Samples were transferred from -80 °C to an incubation at 100 °C for 10 minutes, supplemented with 500 µL of 1x TE, vortexed for 15 seconds at maximum speed, and further incubated at 100 °C for 10 minutes. Premeasured 150 mg of 0.1 mm glass disruption beads (Research Products International 9830, Mt. Prospect, IL, USA) were added to each tube, and samples were bead-beaten horizontally for 20 minutes at maximum speed on a vortex adapter (QIAGEN, 13000-V1-24, Hilden, Germany). Lysates were transferred to 1.5-mL LoBind tubes and SSMaC and qPCR proceeded per Olson *et al.*, 2025 (2), including positive and negative extraction controls. The export threshold for the combined FAM channel (targeting IS*6110* and IS*1081*) used in this analysis was 2,250 Δ Rn. If a negative extraction control amplified (<40 Cq), the entire sample set was rerun from reserved eluate with fresh qPCR reagents after decontaminating workspaces and equipment. Sets in which the contamination issue resolved on rerun were retained for analyses; those that did not were excluded.

**Supplemental Tables and Figures**

**Table S1.** List of RePORT South Africa study team members.

| Name | Affiliation |
|---|---|
| Pattamukkil Abraham | Perinatal HIV Research Unit, University of the Witwatersrand |
| Thakiera Allie | South African Tuberculosis Vaccine Initiative, University of Cape Town |
| Cynthia Baard | Department of Paediatrics and Child Health, University of Cape Town |
| Zainab Baig | Africa Health Research Institute |
| John Belisle | Colorado State University |
| Nicole Bilek | South African Tuberculosis Vaccine Initiative, University of Cape Town |
| ACE Carstens | Department of Biomedical Sciences, Stellenbosch University |
| Novel N. Chegou | Department of Biomedical Sciences, Stellenbosch University |
| Kevyna Chetty | Africa Health Research Institute |
| Yolundi Cloete | South African Tuberculosis Vaccine Initiative, University of Cape Town |
| Marwou de Kock | South African Tuberculosis Vaccine Initiative, University of Cape Town |
| Gareta Dickman | Africa Health Research Institute |
| Charity Dire | Perinatal HIV Research Unit, University of the Witwatersrand |
| Karen Dobos | Colorado State University |
| Stephany Norah Duda | Vanderbilt Tuberculosis Center, Vanderbilt University Medical Center |
| Mzwandile Erasmus | South African Tuberculosis Vaccine Initiative, University of Cape Town |
| Marina Cruvinel Figueiredo | Vanderbilt Tuberculosis Center, Vanderbilt University Medical Center |
| Marika Flinn | Department of Biomedical Sciences, Stellenbosch University |
| Travis Harris | Vanderbilt Tuberculosis Center, Vanderbilt University Medical Center |
| Andriette Hiemstra | Department of Biomedical Sciences, Stellenbosch University |
| Shameem Jaumdally | University of Cape Town Lung Institute |
| Ryan Johnson | University of Cape Town |
| Farina Karim | Africa Health Research Institute |
| Masooda Kaskar | South African Tuberculosis Vaccine Initiative, University of Cape Town |
| Nobulumko Khomba | South African Tuberculosis Vaccine Initiative, University of Cape Town |
| Thandeka Khoza | Africa Health Research Institute |
| Léanie Kleynhans | Department of Biomedical Sciences, Stellenbosch University; Mater Research Institute – The University of Queensland |
| Tahira Kootbodien | University of Cape Town |
| Andrea Kotze | University of Cape Town Lung Institute |
| Belinda A Kriel | Department of Biomedical Sciences, Stellenbosch University |
| Ané Kruger | Department of Biomedical Sciences, Stellenbosch University |
| Lorraine Lichakane | Perinatal HIV Research Unit, University of the Witwatersrand |
| Ilze Louw | Department of Biomedical Sciences, Stellenbosch University |
| Angelique Luabeya | South African Tuberculosis Vaccine Initiative, University of Cape Town |
| Candice MacDonald | Department of Biomedical Sciences, Stellenbosch University |
| Lindiwe Madziwa | Africa Health Research Institute |
| Lebohang Makhethe | South African Tuberculosis Vaccine Initiative, University of Cape Town |
| Sandisiwe Mangali | South African Tuberculosis Vaccine Initiative, University of Cape Town |
| Linda Mbuthini | University of Cape Town |

| | |
|---|---|
| Carolina Mehaffy | Colorado State University |
| Thabang Moloja | Perinatal HIV Research Unit, University of the Witwatersrand |
| Angelique Mouton | South African Tuberculosis Vaccine Initiative, University of Cape Town |
| Evans Muchiri | South African Tuberculosis Vaccine Initiative, University of Cape Town |
| Mbusiseni Ngema | Perinatal HIV Research Unit, University of the Witwatersrand |
| Hlengiwe Nkambule | South African Tuberculosis Vaccine Initiative, University of Cape Town |
| Onke Nombida | South African Tuberculosis Vaccine Initiative, University of Cape Town |
| Sarah Nyangu | South African Tuberculosis Vaccine Initiative, University of Cape Town |
| Fajwa Opperman | South African Tuberculosis Vaccine Initiative, University of Cape Town |
| Gregory Ording-Jespersen | Africa Health Research Institute |
| Kennedy Otwombe | Perinatal HIV Research Unit, University of the Witwatersrand |
| Will Ramsay | Vanderbilt Tuberculosis Center, Vanderbilt University Medical Center |
| Tracy Richardson | Department of Biomedical Sciences, Stellenbosch University |
| Carmen Segelaar | South African Tuberculosis Vaccine Initiative, University of Cape Town |
| Jane Shaw | Department of Biomedical Sciences, Stellenbosch University |
| Kimberly Shelton | Colorado State University |
| Justin Shenje | South African Tuberculosis Vaccine Initiative, University of Cape Town |
| Theresa Smit | Africa Health Research Institute |
| Bronwyn Smith | Department of Biomedical Sciences, Stellenbosch University |
| Candice Snyders | Department of Biomedical Sciences, Stellenbosch University |
| Marcia Steyn | South African Tuberculosis Vaccine Initiative, University of Cape Town |
| Sara Suliman | University of California San Francisco |
| Floris Swanepoel | Perinatal HIV Research Unit, University of the Witwatersrand |
| Lorraine Thobakgale | Perinatal HIV Research Unit, University of the Witwatersrand |
| Susanne Tonsing | Department of Biomedical Sciences, Stellenbosch University |
| Nicolette Tredoux | South African Tuberculosis Vaccine Initiative, University of Cape Town |
| Megan Turner | Vanderbilt Tuberculosis Center, Vanderbilt University Medical Center |
| Petrus Tyambetyu | South African Tuberculosis Vaccine Initiative, University of Cape Town |
| Habibullah Valley | South African Tuberculosis Vaccine Initiative, University of Cape Town |
| Lyle van de Berg | University of Pretoria |
| Yuri van der Heijden | Vanderbilt Tuberculosis Center, Vanderbilt University Medical Center |
| Gian van der Spuy | Department of Biomedical Sciences, Stellenbosch University |
| Ilana R van Rensburg | Department of Biomedical Sciences, Stellenbosch University |
| Johanna E van Rooyen | South African Tuberculosis Vaccine Initiative, University of Cape Town |
| Hilary Vansell Riley | Vanderbilt Tuberculosis Center, Vanderbilt University Medical Center |
| Ashley Veldsman | South African Tuberculosis Vaccine Initiative, University of Cape Town |
| Lindsay Wilson | University of Cape Town |
| Lesley Workman | Department of Paediatrics and Child Health, University of Cape Town |
| Heather Zar | Department of Paediatrics and Child Health, University of Cape Town |

**Table S2.** Demographic and clinical characteristics of participants in the Clinic Cohort, differentiated by testing method.

| | **Total participants (N = 673)** | **High-volume Method (N = 334)** | **SSMaC Method (N = 339)** | ***p*-value (High-volume vs SSMaC)** |
|---|---|---|---|---|
| | **n (%)** | **n (%)** | **n (%)** | |
| Site | | | | |
| *AHRI* | 312 (46.4) | 153 (45.8) | 159 (46.9) | 0.77 |
| *PHRU* | 28 (4.2) | 16 (4.8) | 12 (3.5) | 0.40 |
| *SUN* | 103 (15.3) | 47 (14.1) | 56 (16.5) | 0.39 |
| *UCT-LI* | 172 (25.6) | 85 (25.4) | 87 (25.7) | 0.93 |
| *UP* | 58 (8.6) | 33 (9.9) | 25 (7.4) | 0.25 |
| Age, years (median, IQR) | 36 (28–44) | 36 (27–44) | 36 (28–45) | N/A |
| Sex (female) | 245 (36.4) | 115 (34.4) | 130 (38.3) | 0.29 |
| HIV Positive | 204 (30.3) | 99 (29.6) | 105 (31.0) | 0.69 |
| Any symptom | 673 (100) | 334 (100) | 339 (100) | 0.99 |
| *Cough* | 588 (87.4) | 293 (87.7) | 295 (87.0) | 0.79 |
| *Fever* | 215 (31.9) | 108 (32.3) | 107 (31.6) | 0.85 |
| *Weight loss* | 365 (54.2) | 191 (57.2) | 174 (51.3) | 0.12 |
| *Fatigue* | 323 (48.0) | 163 (48.8) | 160 (47.2) | 0.67 |
| *Night sweats* | 370 (55.0) | 190 (56.9) | 180 (53.1) | 0.32 |
| *Chest pains* | 258 (38.3) | 140 (41.9) | 118 (34.8) | 0.06 |
| Smoking | | | | |
| *Unknown* | 1 | 0 | 1 | 0.32 |
| *Current* | 299/672 (44.4) | 157 (47.0) | 142/338 (42.0) | 0.19 |
| *Former* | 77/672 (11.4) | 34 (10.2) | 43/338 (12.7) | 0.31 |
| *Never* | 296/672 (44.0) | 143 (42.8) | 153/338 (45.3) | 0.52 |
| Prior TB | 148 (22.0) | 73 (21.9) | 75 (22.1) | 0.95 |
| Chest radiograph | | | | |
| *Unknown* | 332 | 167 | 165 | 0.73 |
| *Normal* | 176/341 (51.6) | 86/167 (51.5) | 90/174 (51.7) | 0.97 |
| *Abnormal* | 165/341 (48.4) | 81/167 (48.5) | 84/174 (48.3) | 0.97 |
| Positive sputum smear microscopy | 165 (24.5) | 79 (23.7) | 86 (25.4) | 0.61 |
| *Not done* | 4 | 2 | 2 | 1.0 |
| *3+ grade* | 45/165 (27.3) | 17/79 (21.5) | 28/86 (32.6) | 0.11 |
| *2+ grade* | 53/165 (32.1) | 31/79 (39.2) | 22/86 (25.6) | 0.06 |
| *1+ grade* | 38/165 (23.0) | 17/79 (21.5) | 21/86 (24.4) | 0.66 |
| *Scanty grade* | 28/165 (17.0) | 14/79 (17.7) | 14/86 (16.3) | 0.81 |
| Positive sputum culture | 206/672 (30.7) | 100/330 (30.3) | 106/338 (31.4) | 0.76 |

| | | | | |
|---|---|---|---|---|
| *Not done* | 5 | 4 | 1 | 0.18 |
| *MGIT time to positivity, days (Median, IQR)* | 9 (6–14) | 10 (6–15) | 9 (6–14) | N/A |
| Positive sputum Xpert Ultra | 221/669 (33.0) | 110 (32.9) | 111/335 (33.1) | 0.95 |
| *Not done* | 4 | 0 | 4 | 0.04* |
| *High SQ grade* | 77/221 (34.8) | 33/110 (30.0) | 44/111 (39.6) | 0.13 |
| *Medium SQ grade* | 42/221 (19.0) | 21/110 (19.1) | 21/111 (18.9) | 0.97 |
| *Low SQ grade* | 56/221 (25.3) | 31/110 (28.2) | 25/111 (22.5) | 0.33 |
| *Very low SQ grade* | 26/221 (11.8) | 15/110 (13.6) | 11/111 (9.9) | 0.40 |
| *Trace SQ grade* | 19/221 (8.6) | 10/110 (9.1) | 9/111 (8.1) | 0.79 |
| *SQ grade not recorded* | 1/221 (0.5) | 0/110 (0.0) | 1/111 (0.9) | 0.32 |

**AHRI**, Africa Health Research Institute. **PHRU**, Perinatal HIV Research Unit, University of the Witwatersrand. **SUN**, Stellenbosch University. **UCT-LI**, University of Cape Town-Lung Institute. **UP,** University of Pretoria. SSMaC, sequence-specific magnetic capture. HHC, household contact. SQ, semi-quantitative. IQR, inter-quartile range. *$p < 0.05$.

**Table S3.** Sensitivity and specificity of tongue swab qPCR in the Clinic and HHC Cohorts; results from **A.** only regular Copan flocked swabs, and **B**. only swabs that are not regular Copan flocked swabs. The *p*-values show whether the sensitivity or specificity of the cohort/method is significant relative to the overall sensitivity or specificity. **p*-value is significant at $p < 0.05$ by 2-population proportion Z-test. SSMaC, sequence-specific magnetic capture. HHC, household contact.

A.

| | **Sensitivity** | | | **Specificity** | | |
|---|---|---|---|---|---|---|
| | **n/N** | **%<br>(95% CI)** | ***p*-value** | **n/N** | **%<br>(95% CI)** | ***p*-value** |
| **Clinic Cohort** | | | | | | |
| High-volume qPCR | 65/102 | 63.7%<br>(53.6–73.0) | 0.99 | 188/200 | 94.0%<br>(89.8–96.9) | 0.87 |
| SSMaC with qPCR | 79/107 | 73.8%<br>(64.5–81.9) | 0.92 | 192/204 | 94.1%<br>(90.0–96.9) | 0.83 |
| **HHC Cohort** | | | | | | |
| High-volume qPCR | 8/27 | 29.6%<br>(13.8–50.2) | 0.70 | 103/106 | 97.2%<br>(92.0–99.4) | 0.08 |

B.

| | **Sensitivity** | | | **Specificity** | | |
|---|---|---|---|---|---|---|
| | **n/N** | **%<br>(95% CI)** | ***p*-value** | **n/N** | **%<br>(95% CI)** | ***p*-value** |
| **Clinic Cohort** | | | | | | |
| High-volume qPCR | 2/3 | 66.7%<br>(9.4–99.2) | 0.92 | 13/13 | 100%<br>(75.3–100) | 0.38 |
| SSMaC with qPCR | 3/5 | 60.0%<br>(14.7–94.7) | 0.52 | 19/19 | 100%<br>(82.4-100) | 0.30 |
| **HHC Cohort** | | | | | | |
| High-volume qPCR | 7/17 | 41.2%<br>(18.4–67.1) | 0.60 | 22/30 | 73.3%<br>(54.1–87.7 | 0.004* |

**Figure S1.** Tongue swab, sputum culture and smear, chest radiograph, and symptom positivity (%) by sputum Xpert Ultra grade among **(A)** symptomatic and **(B)** asymptomatic participants. The numbers within the bars are the test result N. HHC, household contact. TS, tongue swab. CXR, chest radiograph.

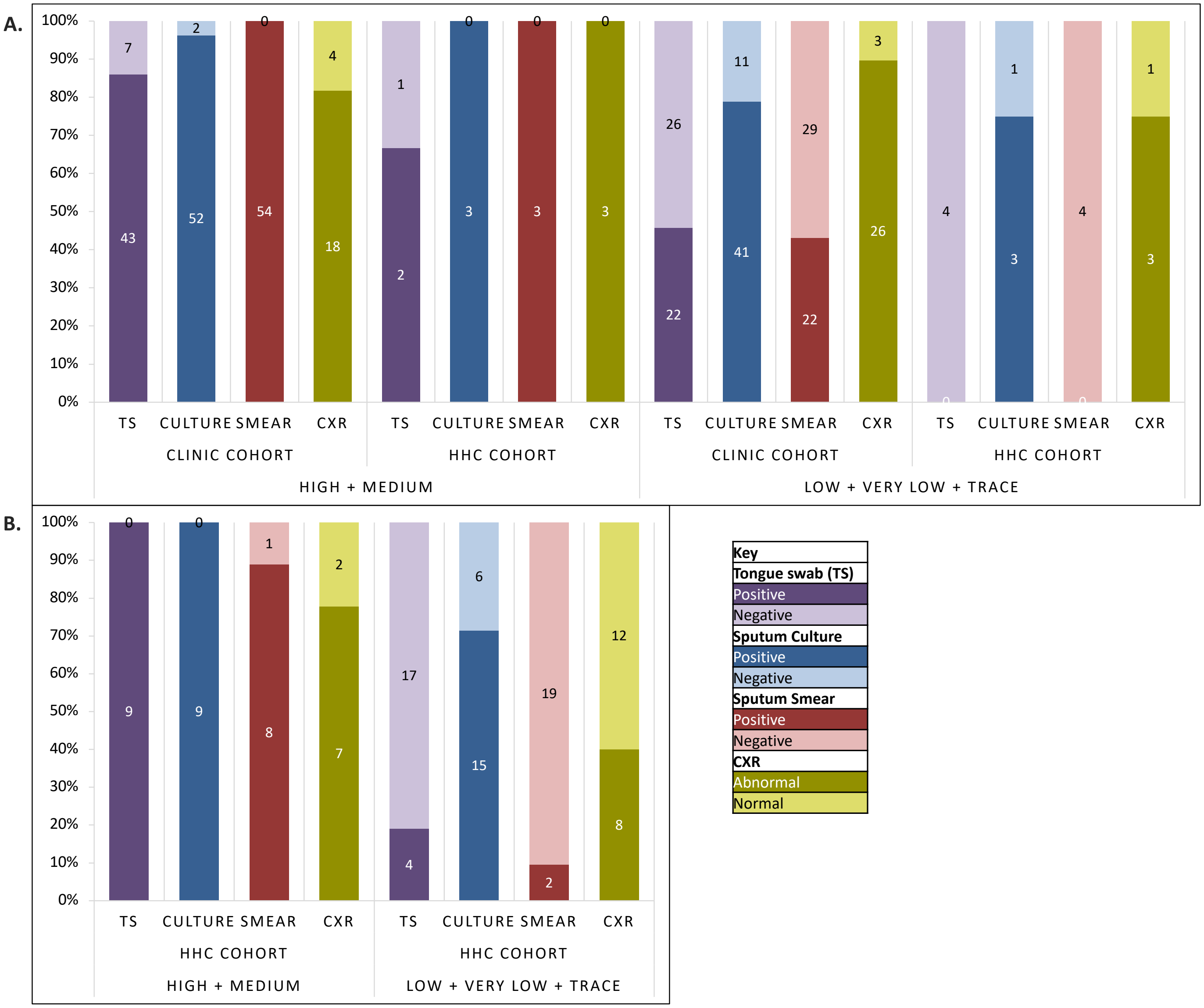

**Figure S2.** Proportion of sputum Xpert Ultra semi-quantitative grades in TB Cases by cohort. Sputum Xpert Ultra semi-quantitative results are provided as a proxy for MTB bacillary load. Cx, culture. HHC, household contact.

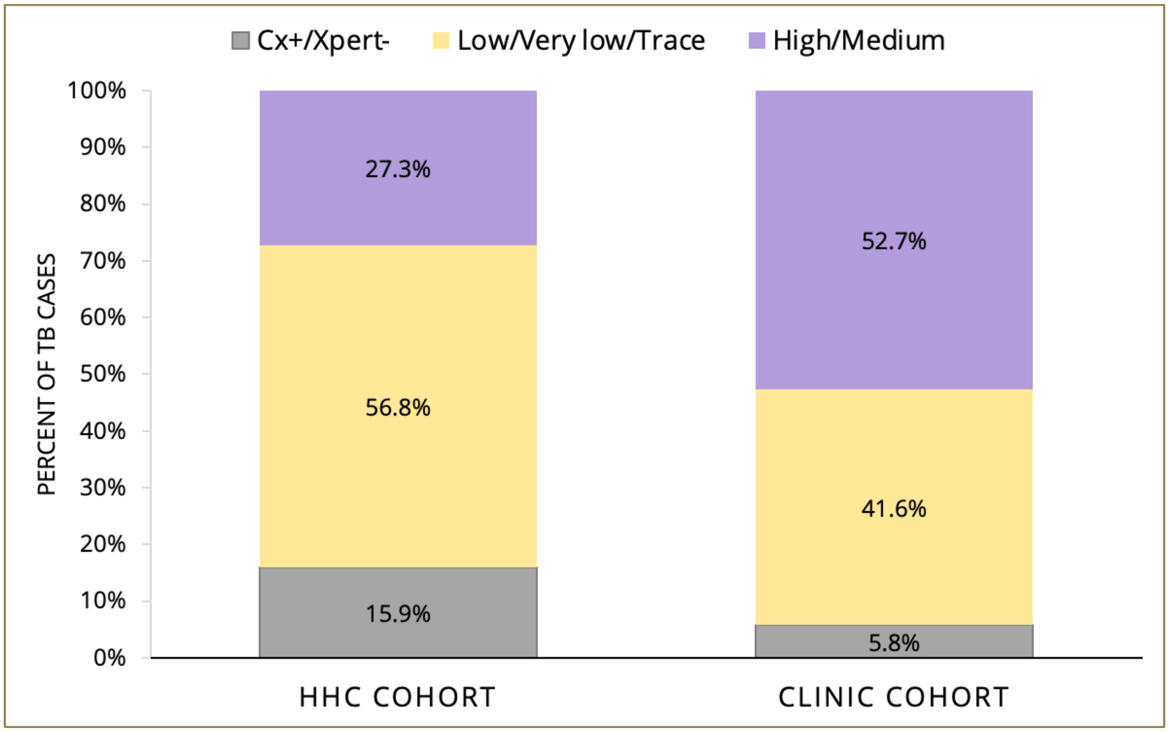
